# A novel model to quantify blood transit time in cerebral arteries using ASL-based 4D magnetic resonance angiography with example clinical application in moyamoya disease

**DOI:** 10.1101/2024.10.09.24315081

**Authors:** Alex A. Bhogal, Simone M. Uniken-Venema, Pieter Deckers, Maarten Versluis, Kees P. Braun, Albert van der Zwan, Jeroen C.W. Siero

## Abstract

Angiography is critical for visualizing cerebral blood flow in intracranial steno-occlusive diseases. Current 4D magnetic resonance angiography (MRA) techniques primarily focus on macrovascular structures, yet few have quantified hemodynamic timing. This study introduces a novel model to estimate macrovascular arterial transit time (mATT) derived from arterial spin labeling (ASL)-based 4D-MRA. We provide examples of our method that visualize mATT differences in patients with intracranial steno-occlusive disease (moyamoya and atherosclerosis) and changes in mATT resulting from the cerebrovascular reactivity response to an acetazolamide (ACZ) injection. Furthermore, we present a method that projects sparse arterial signals into a 3D native brain-region atlas space and correlate regional mATT with other hemodynamic parameters of interest such as tissue transit time and cerebrovascular reactivity. This approach offers a non-invasive, quantitative assessment of macrovascular dynamics, with potential to enhance understanding of large-vessel and tissue-level hemodynamics and augment monitoring of treatment outcomes in steno-occlusive disease patients. Furthermore, it sets the stage for more in-depth investigations of the macrovascular contribution to brain hemodynamics.

## Introduction

Angiography has been a key technique for visualizing blood flow in the large (≥2-3 mm) and medium (1-2 mm) cerebral arteries, such as the middle cerebral and basilar arteries, along with their branches. Information on blood flow characteristics provides critical insights for diagnosing and staging neurovascular disease^1^, especially for steno-occlusive conditions such as moyamoya, arteriovenous malformations and aneurysms^2^ or moyamoya vasculopathy.^3^ This understanding can aid decision-making regarding the need for bypass surgery and planning endovascular procedures.

Digital subtraction angiography (DSA) provides the highest spatial resolution in a time-resolved manner, and is capable of resolving even smaller arteries/arterioles (<1 mm). However, DSA required catheterization (carrying a risk of neurologic complications)^4^, uses a contrast agent and involves exposure to radiation.^5^ For these reasons, routine clinical evaluation or cerebral vessel architecture is done using computed tomography angiography (CTA), which also requires intravenous injection of contrast (no catherization) and exposure to ionizing radiation, time-of-flight (TOF) or contrast-enhanced (CE) magnetic resonance imaging (MRI). While free of radiation and non-invasive in nature, these techniques do not provide dynamic information that might indicate blood arrival timing, flow or velocity.

The past decade has seen the emergence of arterial-spin-labelling (ASL) based 4D-MRA techniques as alternatives to visualize (pathological) vessel structure. These methods have gained interest as a “best of both worlds” technique, allowing non-invasive and contrast-free intracranial magnetic resonance angiography (MRA) with three-dimensional (3D) spatial and dynamic acquisition (4D).^6^ Like DSA, 4D-MRA can be used to selectively target vessels, and can visualize collateral flow via the circle of willis^7^, but also at the level of distal arteries and leptomeningeal collateral vessels.^8^ More recently, 4D ASL-based MRA has been combined with super-selective labelling of individual arteries to visualize individual territorial flow.^9, 10^ This innovation has allowed the assessment of feeding arteries in intracranial arteriovenous malformations (AVM) and dural arteriovenous fistulas (AVF).^11^

Moreover, vessel-selective 4D ASL-based MRA showed the potential to better evaluate bypass patency and intracranial collaterals after bypass surgery than traditional 3D time-of-flight (TOF) angiography.^12^ Thorough reviews of different 4D-MRA techniques have been presented by Suzuki et al. (2019)^13^ and Togao et al, (2023).^14^

Thus far, 4D-MRA techniques have focused on visualizing and understanding arterial blood flow patterns and their relation to pathology, with limited reports focusing on quantifying associated hemodynamic timing.^6,15^ Knowledge of blood arrival times throughout the macrovasculature might inform on the cerebral hemodynamic status, including the presence and efficiency of collateral circulation,^16, 17^ which may be an important prognostic marker in patients with moyamoya vasculopathy as well as acute ischemic stroke.^18^ While the arterial transit time (ATT)^19, 20^ of labelled blood water in the tissue can be calculated using ASL with multiple post-labeling delays (multi-PLD ASL), the ATT metric may be insensitive in distinguishing slow-flowing blood resulting from a drop in perfusion pressure associated with blood arriving through longer collateral pathways. This leads to longer transit times, that are reflected in the ATT parameter, which is measured distally at the tissue level. Therefore, this study aims to present a new model that utilizes 4D ASL-based MRA data to estimate the macrovascular transit time (mATT) of labeled blood water in the large conduit vessels, including the internal carotid (ICA), middle cerebral (MCA), and anterior communicating arteries (ACA), along with their branches. We provide examples of our method that visualize mATT differences in patients with Moyamoya disease and changes in mATT resulting from the cerebrovascular reactivity response to an acetazolamide (ACZ) injection.

## Methods

### Study population

The Medical Ethics Review Committee of the University Medical Centre Utrecht declared that the Medical Research Involving Human Subjects Act (WMO) did not apply to the present research since all study measures were part of routine clinical practice. All patients or their legal representative

(i.e., parent or guardian) provided written informed consent to use their data. The study population consisted of patients with intracranial steno-occlusive disease, irrespective of the underlying cause (moyamoya, atherosclerosis), with a clinical indication to undergo hemodynamic imaging with an ACZ challenge to measure cerebrovascular reactivity (CVR). In this protocol, cerebrovascular reactivity (CVR) is measured via an acetazolamide (ACZ) injection using both multi-PLD ASL, acquired pre– and post-ACZ. In short, the ACZ injection was performed 60 seconds after the pre-ACZ multi-PLD ASL scan using a dose of 20 mg/kg (maximum 1 g) in 30 cc of 0.9% NaCl solution and a flowrate of ∼0.3 cc/s. The post-ACZ multi-PLD ASL was started 15 min after the ACZ injection. The scan indications varied, but the most common were to assess the need for bypass surgery, to re-evaluate patients in whom surgery was previously not deemed necessary but who developed new ischemic symptoms, or for hemodynamic evaluation after neurosurgical intervention. For full protocol details, see Deckers et al., 2023.^21^

After retrospectively surveying our database, twelve distinct patients were identified in which 4D-MRA data were acquired after ACZ injection, and informed consent was provided (see supplementary table 1 for patient data). Two patients were excluded from analysis due to motion-related artifacts, leaving ten post-ACZ scans. In five of twelve patients, 4D-MRA data were acquired both pre– and approximately five minutes post-ACZ injection. One of the five post-ACZ scans was excluded due to motion, leaving four pre– and post-ACZ scans for comparison. It should be noted that the remaining 6 post-ACZ scans were obtained approximately 15 minutes post-injection. Data were also acquired in 2 healthy subjects under a sequence development ethical protocol, which was approved by the Medical Ethics Review Committee of the University Medical Centre Utrecht. These subjects only received a baseline 4D-MRA without an additional ACZ challenge. Informed consent was given by each healthy subject.

### MRI Data Acquisition

Data was acquired using a 3T scanner (Philips Healthcare, Best, The Netherlands) and a 32-channel receive coil (Nova Medical Inc.). A vendor-specific (Philips) four-dimensional dynamic magnetic resonance angiography (4D-MRA) sequence using a pseudo-continuous arterial spin labelling (pCASL) scheme was used^6^. The Philips implementation also takes advantage of ‘keyhole’ and ‘view-sharing’ techniques as part of the 4D-PACK implementation for scan acceleration. Full sequence details are available in Obaro et al., 2018^6^. Our parameters were as follows: 4D MRA using a 3D GRE readout, 8 time-points at 100, 200, 400, 600, 800, 1200, 1600 and 2200⍰ms, 75% keyhole central size, turbo factor=60, SENSE factor=3×1.5 (right-left, feet-head direction), half scan (partial Fourier) factor = 0.8×0.8 (right-left, feet-head direction), TR/TE/flip angle =⍰6⍰ms/1.97⍰ms/11°, acquired voxel size 1×1.4×1.6⍰mm^3^, reconstructed voxel size 0.6×0.6×0.8 mm^3^, field-of-view = 200×200×120 mm^3^, scan time: 6⍰min 16⍰s).

For the multi-PLD ASL measurements, a dynamic pCASL sequence with a variable repetition time scheme (vTR-ASL) and optimized background suppression was used, see for more details Togao et al.^22^ In short, the vTR-ASL sequence allows arbitrary combinations of label durations and post-labelling delays, rendering it a time-efficient multi-PLD ASL method to obtain CBF and arterial transit time (ATT) measurements. Improved motion-sensitized driven-equilibrium (iMSDE) was used for arterial signal suppression.^23^ Our parameters were as follows: pCASL using a 3D GRASE readout, 10 label durations at 300, 800, 1300, 1800, 2000, 2000, 2000, 2000, 2000, 2000⍰ms, 10 post-labelling durations at 200, 200, 200, 200, 500, 1000, 1500, 2000, 2500, 3000, 4 background suppression pulses, iMSDE TE=16ms, iMSDE velocity encoding = 5 cm/s in all 3 directions, turbo spin echo factor = 15, EPI factor= 15, SENSE factor=2.5×1 (anterior-posterior, feet-head direction), half scan (partial Fourier) factor = 1×0.8 (right-left, feet-head direction), TR/TE =⍰variable (1650-6150 ms) /15⍰ms, excitation/refocusing flip angle = 90°/180°, acquired and reconstructed voxel size = 4×4×8⍰mm^3^, field-of-view = 256×240×120 mm^3^, scan time: 2⍰min 54⍰s). An equilibrium magnetization (M0) calibration image for CBF quantification was acquired with a repetition time of 3000 ms and identical imaging parameters.

V-TR ASL CBF and ATT quantification was performed using the general kinetic model of Buxton by applying nonlinear fitting^24^ and following fixed fit parameters: labelling efficiency α_label_=0.85, background suppression efficiency α_BGS_=0.8, T1t (tissue T1)=1.5 s, T1b (arterial blood T1)= 1.66 s, tissue-blood partition coefficient λ=0.9. ASL-CVR was computed by subtracting the pre-ACZ map from the post-ACZ CBF map after rigid body co-registration, yielding CVR in ΔCBF in units ml/100gr/min.

### Pre-processing and mATT Fitting

Data were processed using in-house developed MATLAB scripts (MATLAB 2023b, The Mathworks Inc., Natick, Massachusetts). Difference images were generated via pair-wise subtraction of label and control pairs for each time-point separately. Next, a series of filtering and masking steps were performed to isolate vascular structures.

First, a brain mask was generated based on the first time-point 4D-MRA magnitude inflow image (FSL: BET^25^). This mask was modified in order to remove artifacts located around the eyes without extracting extracranial vessels that were also labelled. Next, a 2D Frangi filter^26^ was applied in all three cartesian dimensions. The resulting images were binarized to create vessel masks for each filter response direction. A final vessel mask was created consisting of only the voxels that were common in each directional mask. An area opening process was then applied using the MATLAB function ‘*bwareopen.m*’. This function was tuned on a per-subject basis, to remove ‘blob-like’ structures arising from high tissue pulsatility around the base of the brain around the circle of Willis and large cerebral vessels. Finally, an edge-preserving bilateral smoothing filter was applied using the final vessel mask as the guide image. Here, a full-width-half-maximum (FWMH) of 5×5×4 voxels corresponding to a 2.98×2.98×3.20 mm filter kernel size was used. This filter size was chosen to approximate the diameter of the larger intracranial vessels^27^. Finally, to estimate mATT, we used a piece-wise saturation signal model to describe the observed signal increase with respect to time (see figure 1 and supplement for MATLAB code).

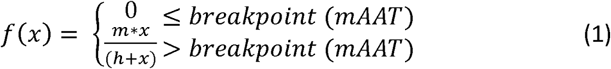

**Figure 1.**
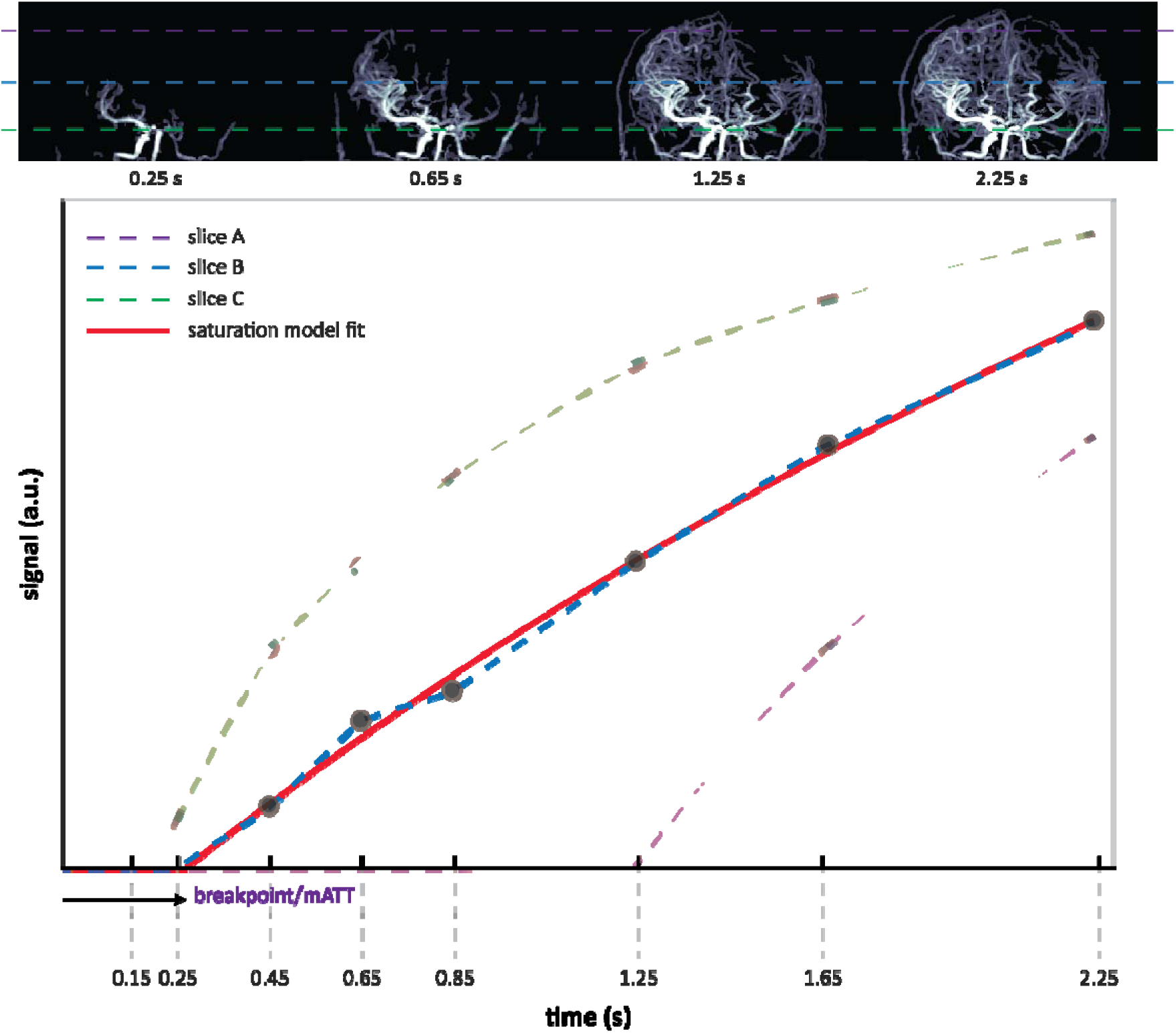
-top: 4D-MRA maximum intensity projections show inflowing labelled spins at a selection of time-points. The dashed lines represent individual slices; 1-bottom: the mean signal for each slice is plotted and the data, as a function of the acquisition time, is fit to the two-piece macrovascular arrival time model (equation 1). Slices (or individual voxels) located more cranially receive labelled blood at later time-points. This, and the assumption that the signal evolution will saturate over time, allow the extrapolation of the macrovascular transit time (mATT) per voxel.

The function uses two pieces: an initial linear period (y = 0) represents the time taken for labelled spins to arrive at the voxel of interest. After a breakpoint, the second piece is described by a nonlinear saturation function (equation 1), which describes the progressive increase in the number of labelled spins occupying a given voxel. Here, ‘m’ represents the saturation level or maximum signal that f(x) approaches at late acquisition times, and ‘h’ represents the time at which f(x) is at half maximum (i.e. ½*m). We interpret the breakpoint as the mATT.

### Projection of mATT to Native Atlas Space

After fitting mATT, the first time-point magnitude image from the 4D-MRA scan was registered to the ICBM 2009c nonlinear asymmetric 1mm MNI template^28^ using affine and nonlinear transformations using elastix^29^ (MATLAB wrapper implementation available in the seeVR toolbox^30^). Transformations were then inverted and applied to the MNI-registered Allen Brain Atlas, which delineates 282 distinct brain regions^31^. Volumetric mATT information was then translated to the native atlas space by calculating the mean mATT of all arterial voxels contained within a given atlas regions. This average mATT was then assigned to that entire region including voxels in which no information was present in mATT maps (see figure 2). Parameter maps of interest, such as ATT maps derived using a variable TR pCASL sequence used within our clinical protocol, were also registered to the first time-point magnitude image (and therefore also the native atlas). This allowed the averaging of additional hemodynamic information within the same regions of interest, thus facilitating correlation analysis.

**Figure 2A:**
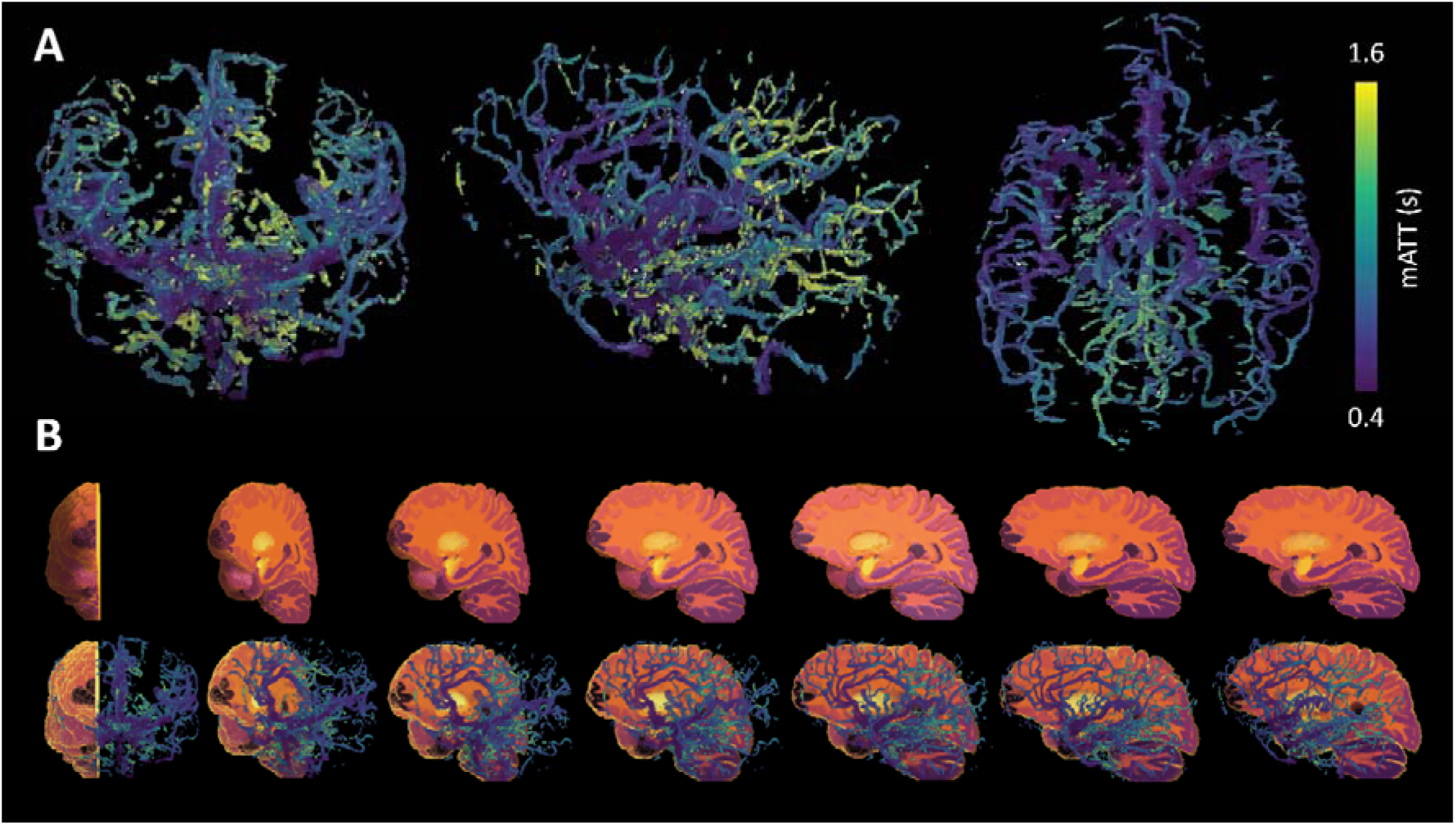
Coronal, sagittal and transverse projections of the volumetric mATT visualization are shown for a healthy subject; 2B: top row shows a cut-out volumetric section of the Allen Brain Atlas where differences in colours delineate specific regions of interest. The mATT maps are overlaid on the atlas, allowing the average mATT for all voxels within a specific region to be calculated. This regional mATT value is then assigned to the entire region of interest to produce parametric maps of mATT (shown in figures 4-6). Note the expected increase in mATT from anterior to posterior brain regions and in the direction of the bottom to the top of the brain (caudal to cranial).

### Statistical analysis

In addition to the whole-brain mean and standard deviation mATT for each subgroup (healthy vs. diseased, pre-vs. post-ACZ), the mean and standard deviation of the mATT was calculated as a function of distance from the first imaging slice in the caudal to cranial direction). With this approach we ignore the effect of pathology and assume that the mATT will mainly change as a function of the distance from the major input arteries (ICA < MCA < ACA < branches). Linear regression was performed on the mean value curves to estimate the increase in mATT as a function of distance (slope). The adjusted r-squared values are reported as a measure of the goodness of fit. In a single case, linear regression analysis was performed using ROI-averaged (i.e. native atlas regions) mATT and ATT values derived from 4D-MRA and vTR pCASL scans, respectively.

### Results and Cases

The global maximum mATT (mATT_tot_) is an indicator of total time taken to reach the most visible distal vessels (>1mm). The mean +/− std mATT_tot_ values for pre– and post-ACZ groups (n = 4) were 2148 +/− 58 ms and 2205 +/− 31 ms, respectively. In the healthy subject cases, the average mATT_tot_ was 2247 ms (range 2247-2248ms), while in the total patient group, average mATT_tot_ was 2142 +/− 173 ms. The range of mATT_tot_ across all 4 groups was 105 ms, suggesting that proximal vessels with similar transit times were identified in all groups and that the ability to resolve later arriving blood may be limited by the currect sequence parameters.

To assess the spatial dependence of mATT, linear regression was performed between the average slice-wise mATT and distance when moving from caudal to cranial slices (i.e. distance from labelling plane: see figure 3). When comparing the pre-and post-ACZ patients (n = 4), the ACZ effect after five minutes was sufficient to reduce the slope of the regression by 1.1 ms/mm – going from 7.7 ms/mm (adjusted R-squared = 0.96) to 6.6 ms/mm (adjusted R-squared = 0.92). In healthy subjects (n = 2), the mean slope was 5.1 ms/mm (adjusted R-squared = 0.84) at baseline, while in the total patient group (n = 10) the slope was 5.7 ms/mm (adjusted R-squared = 0.96) post-ACZ. Although we do not have sufficient healthy subject data to perform statistical analysis, it would appear that the effect of ACZ in the patient group reduced mATT towards a level that could be expected in healthy subjects.

**Figure 3:**
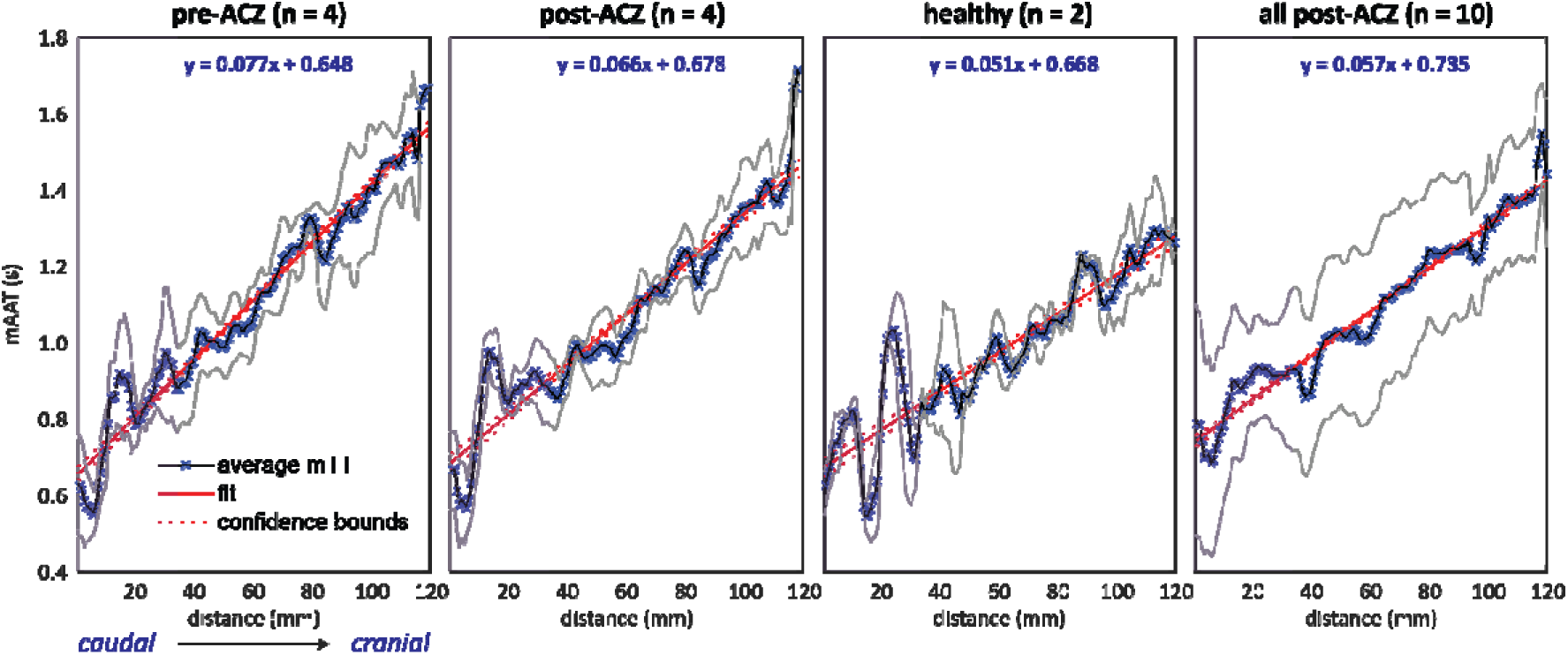
Average mATT calculated as a function of the distance from the first imaging slice (caudal to cranial) for four subgroups. Black lines (marked with ‘x’) represent the average mATT value, while gray shaded areas represents the standard deviation at each distance. The purple shaded region denotes an area of interest in oscillating mATT values may represent arrival of blood through the carotid syphon or possible flow-related artifacts that appeat common across subjects. Increased slopes and maximum mATT at cranial locations for the patient data compared to the healthy subjects demonstrate the increased overall arterial transit times associated with steno-occlusive disease.

Based on this linear model, the average mATT at 120 mm, approximately the distance from the labelling plane to the most distal visible vessels using the current acquisition parameters, for the pre-ACZ, post-ACZ, healthy and all post-ACZ were 1572, 1470, 1280 and 1419 ms, respectively.

We present three examples from our database to highlight the possible added value of our mATT model.

Example 1: A patient in their mid 50’s with bilateral moyamoya disease and no previous surgery who has suffered from transient ischemic attacks in both hemispheres. Data was acquired before and after the administration of ACZ. The maximum intensity of the 4D-MRA inflow images qualitatively showed a clear increase in the visibility of distal vessels upon ACZ injection (figure 4A, supplementary figure 1), and a reduction in the arrival time of the labelled blood, which correlated with stenosis of the left M1 and A1, and was confirmed in the fitted mATT parameter (figure 4B). In both examples, mATT increased when moving from the base (caudal) to the top of brain (cranial), as expected due to the long downstream transit path. Regional changes in mATT could be visualized in the native atlas space. Subtraction of the pre– and post-ACZ mATT maps in atlas space revealed asymmetric bilateral reductions in mATT (figure 4C) on the order of 0.2 s. Notably, there was a greater post-ACZ reduction of mATT in the left (affected) hemisphere compared to the right (unaffected) hemisphere after ACZ. Despite the clear stenosis and delated blood arrival, the left hemisphere showed a good CVR response. Dynamic subtraction angiography images acquired at the referring hospital (see supplementary figure 3) reveal significant contralateral flow that may explain delayed, but adequate baseline perfusion leading to intact CVR. This example highlights the notion that, when combined with a vasodilatory stimulus, the ability of 4D-MRA to visualize distal collateral vessels can be enhanced. Moreover, when combined with additional measures such as CVR, mATT may be an insightful parameter to aid interpretation of complex hemodynamic anomalies.

**Figure 4:**
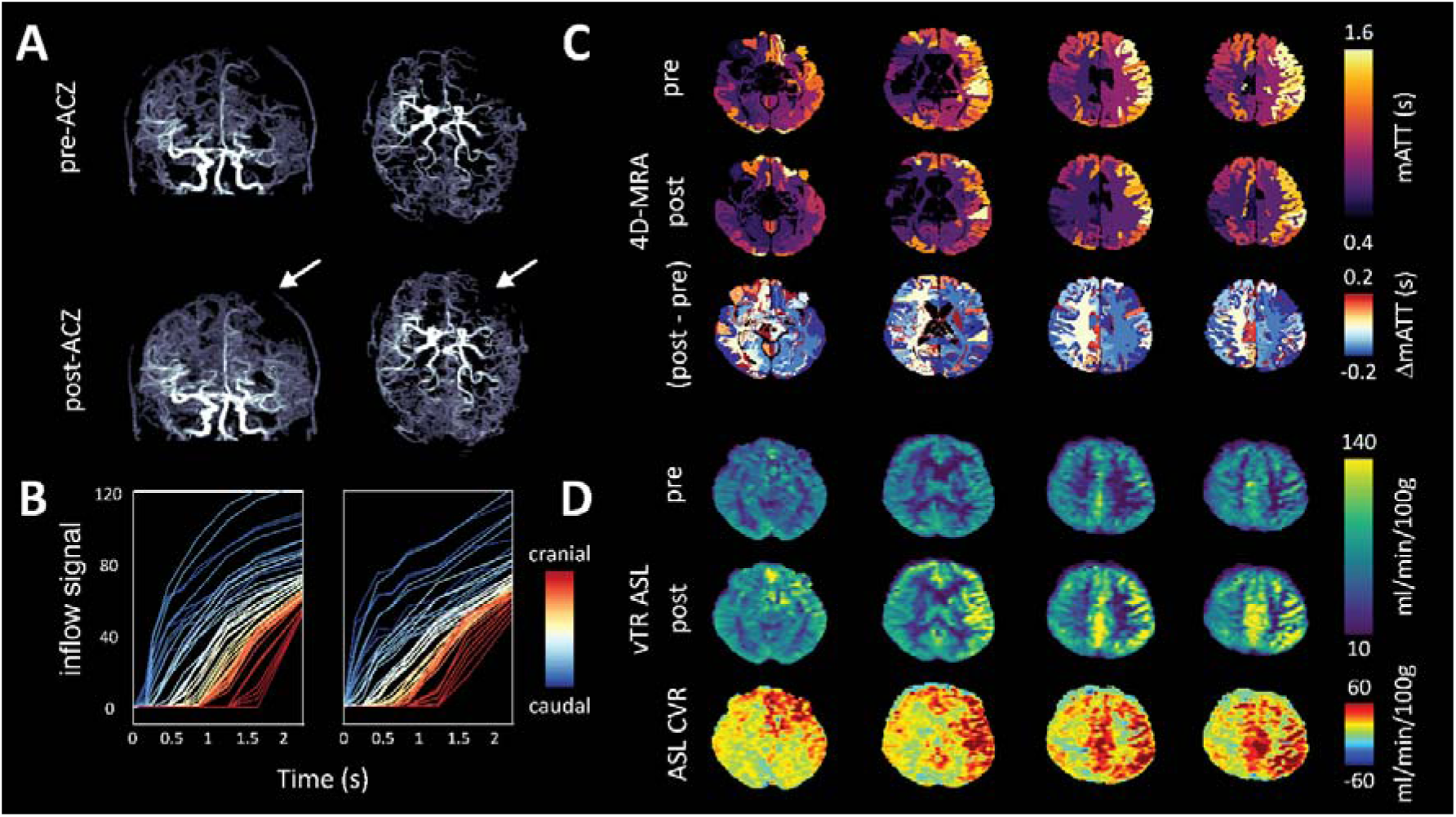
Images of a patient in their mid 50’s with bilateral moyamoya. Angiographically, the left hemisphere (white arrows) is most affected with severe stenosis of the M1 and A1 segment; A: Maximum intensity projections derived from late time point subtraction images (2.25 s) before (top) and approximately 5 minutes after the administration of ACZ; B: average slice-wise (3 slices per curve) inflow signal curves when moving from the base (caudal) to the top (cranial) of the brain. Note that slices located more cranially show increased mATT, as expected. For the associated model-fit curves see figure S-2; C: native atlas representation of mATT before (top) and after (middle) ACZ injection reflect shortened mATT in response to global vasodilation with higher mATT in the affected (right) hemisphere. The subtraction (post-minus pre-ACZ mATT) atlas (bottom) shows the average regional mATT change (maximum on the order of 0.2s). Clear differences can be seen in the response between left and right hemispheres (see discussion for interpretation); D: perfusion images derived from the vTR ASL sequence before (top) and approximately 10 minutes after (middle) ACZ injection show clear perfusion increases in the right hemisphere despite prolonged mATT, which may be explained by delayed arrival due to contralateral flow (see supplementary figure for DSA images).

Example 2: A patient with bilateral moyamoya and left-sided indirect bypass surgery. Native atlas images (figure 5A) show elevated pre-ACZ mATT in the left hemisphere, correlating with less visible large vessel structures in the corresponding inflow 4D-MRA image. After ACZ, mATT values decrease in the left (bypass) hemisphere but show signs of increasing mATT in contralateral regions. This was contrary to expectations, but one possible explanation may be that the contralateral blood supply is augmented by leptomeningeal collaterals to which the 4D-MRA sequence is insensitive. In this patient, a vTR ASL^32^ sequence was also run before and approximately 10 minutes after ACZ injection to acquire CBF and CVR information. Despite the longer working time to evoke the vasodilatory effect (compared to 4D-MRA), the pattern of changes in the arterial transit time (ATT) was maintained at the tissue level (figure 5B). Moreover, regions with prolonged ATT showed reduced CVR or steal phenomenon in the ASL CVR image. Tissue ATT maps derived from the vTR sequence are sensitive to the arrival of blood in the parenchyma, downstream of the larger vessels. As expected, tissue ATT was prolonged when compared with mATT, which is shown by the slopes of the regression lines (i.e. slopes less than 1; figure 5C): 0.38 pre-ACZ (adjusted R-squared = 0.24, p < 0.0001) versus 0.47 post-ACZ (adjusted R-squared = 0.13, p < 0.001). This example highlights the ability to compare mATT times with complementary hemodynamic parameter maps through projection onto the native atlas space. Moreover, macrovascular timing information may suggest the presence of potential collateral flow pathways.

**Figure 5:**
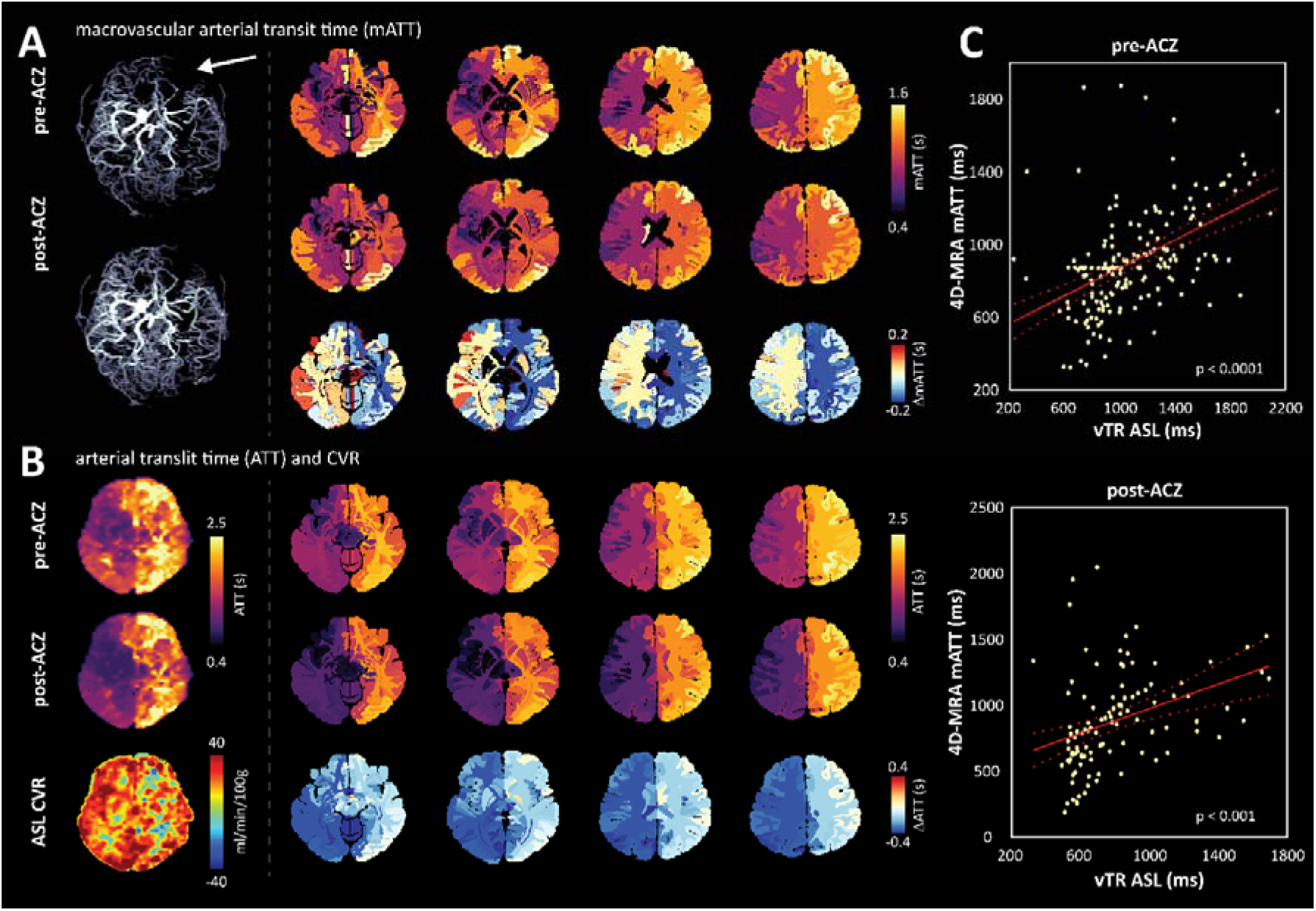
Images of patient with bilateral moyamoya, left-sided indirect bypass; A: Pre– and approximately 5 minutes post-ACZ maximum intensity projections (left) with corresponding mATT atlas images. Pre-ACZ mATT is elevated in the left hemisphere (denoted by white arrow), indicating severe stenosis of the left distal internal carotid artery and M1 segment. The subtraction atlas shows an asymmetric reduction in mATT (right). Interestingly, the ATT reduction is less in the unaffected hemisphere, likely due to the intact CVR response and vessels with longer mATT that become visible after ACZ; 5B: Pre– and approximately 10 minutes post-ACZ multi-PLD ASL ATT maps derived from a vTR multi-PLD ASL sequence. In line with mATT, parenchymal ATT is reduced after ACZ administration; 5C: regression analysis between each modality shows a significant positive correlation between pre– and post-ACZ ATT (vTR ASL) and mATT (4D-MRA).

Example 3: A patient in their mid-30’s with bilateral moyamoya, after a direct extracranial-intracranial bypass on the right after persistent transient ischemic attacks from the right hemisphere. Early blood arrival is evident through the bypass vessel and in the large cerebral vessels in the contralateral hemisphere (figure 6A). The volumetric rendering maps show that the timing through the bypass closely matches the arrival times in the large contralateral cerebral vessels, indicating a successful bypass. These observations are confirmed by the volumetric quantitative mATT data (figure 6B), and also the native atlas representations (figure 6C). This example highlights the potential to evaluate bypass patency and associated blood flow dynamics after treatment using mATT measures from 4DMRA.

**Figure 6:**
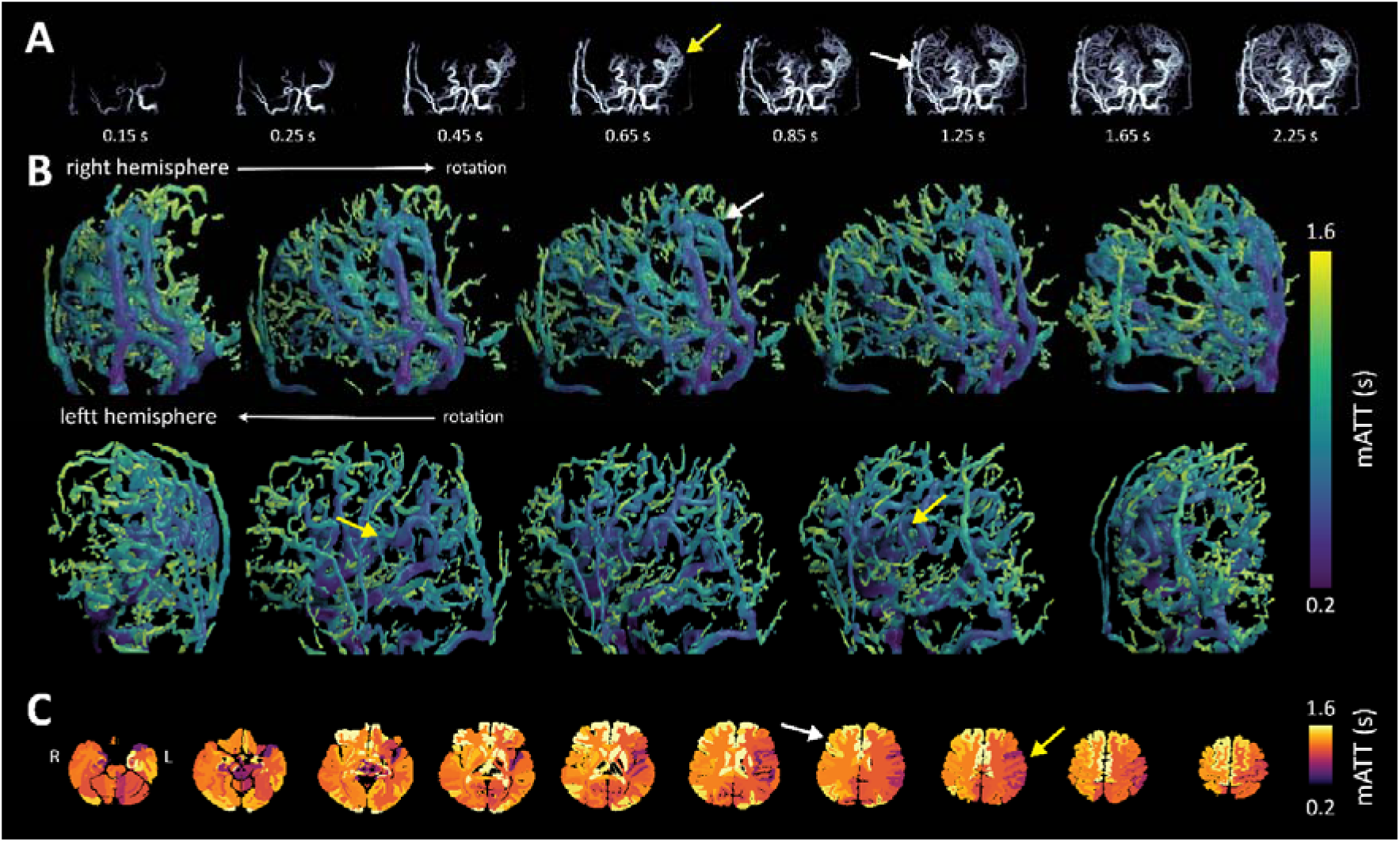
Images of a patient in their mid 30’s patient with bilateral moyamoya, after a direct extracranial-intracranial bypass on the right. A: 4D-MRA images in a moyamoya patient after right-sided bypass. Yellow arrow(s) highlight short mATT in large vessels of the contralateral hemisphere, while white arrow(s) depict delayed arrival through the bypass vessel and subsequently also distal smaller vessels feeding the brain tissue; B: hemispheres confirm the qualitative information provided by the intensity projections; C: atlas mATT images clearly depict regional differences in mATT associated with vascular architecture as highlighted by the yellow (shorter mATT) and white (longer mATT) arrows.

## Discussion

Our study provides three main outcomes: first, we introduce a novel model to quantify signal dynamics from 4D-MRA data by estimating macrovascular arterial arrival times. Second, we developed a processing methodology to translate volumetric arterial measurements into patient-native space, enabling comparison with complementary 3D hemodynamic parameter maps or structural information. Third, we demonstrated the utility of these outcomes by comparing pre– and post-ACZ mATT maps, where we observed the expected decrease in arterial blood arrival time due to vasodilation. Moreover, we can now assess relationships between tissue ATT and CVR obtained by multiPLD ASL. The primary advantages of 4D-MRA are its non-invasive nature, high-resolution imaging, and ability to provide dynamic inflow information of the large cerebral circulation in pathology and health. Our model provides quantitative data to assess the impact of pathology on macrovascular arrival times, identify flow collateralization, and evaluate the efficacy of treatments such as direct bypass.

Our results suggest that using a vasodilatory agent like ACZ can enhance the visibility of distal arteries, which may indicate intact cerebrovascular reactivity (CVR). In four cases with both pre– and post-ACZ data, post-ACZ scans were obtained approximately five minutes after injection. The remaining six post-ACZ datasets were acquired around 15 minutes post-injection, coinciding with the peak vasodilatory effect between 12-15 minutes. This timing suggests that cerebrovascular reactivity was likely sub-maximal in the pre-versus post-ACZ comparison. A global shortening of mean arterial arrival time (mATT) was observed despite this. Affected hemispheres showed higher negative ΔmATT values compared to unaffected ones, with some unaffected regions even displaying positive ΔmATT. This unexpected finding can be attributed to the ACZ challenge, which improves the visibility of distal vessels that have longer mATT, thereby influencing regional mean mATT values. To correct for this, a common vessel mask derived from the pre-ACZ analysis could be applied to both volumetric mATT maps (as shown in supplementary figure 3). Additionally, increasing the degree of parcellation within the atlas template could ensure that mATT values are averaged over smaller regions.

Macrovasular arterial transit time maps generally align with bolus arrival time maps from multi-PLD pCASL used in tissue perfusion mapping,^33^ showing reduced mATT in the anterior and middle cerebral arteries compared to the posterior cerebral artery. Our results, using the longest mATT values to measure mATT_total_, agree with literature values from various contrast techniques. For instance, previous studies reported tissue-level arrival times of 0.9–2.0 seconds using hypoxic arterial BOLD contrast.^34^ Wang et al. (2013) compared multi-PLD ASL with Dynamic Susceptibility Contrast (DSC) and found ATT values of 1.0–3.5 seconds in acute stroke patients, with longer ATT in stroke-affected hemispheres.^35^ The effect of pathology on ATT was supported by Tsujikawa et al. (2016) who observed increased mean ATT for ipsilateral (1.51 ± 0.41 s) versus contralateral territories (1.12 ± 0.30 s) affected by occlusive cerebrovascular disease when using pulsed ASL.

While variability arises from factors like planning differences and structural morphology, any limited visibility of distal vessels in 4D-MRA means we likely underestimate ATT_tot_. These inaccuracies may be mitigated by adding longer label durations. Nevertheless, we are sensitive to changes in mATT, as can be seen by a clear decrease in the regression slope (0.077 to 0.066 s/mm) after ACZ injection (figure 3). We also note a further mATT reduction when looking at the two control subjects (0.051 s/mm) and the complete group of post-ACZ scans (0.054 s/mm), of which 6 were acquired during assumed maximal vasodilation; i.e. 15 minutes post-ACZ rather than 5 minutes post-ACZ as was done in the pre-versus post-injection comparison. The stronger dilatory effect in these 6 subjects likely compensated for the pathology-dependent increase in mATT, leading to values more in line with those seen in the healthy subjects.

### Considerations for data processing

Since most of our example datasets were not influenced by significant head motion, it was not necessary to implement a systematic motion correction between inflow images. However, it is conceivable that motion correction could improve model fitting in some cases. Here, we recommend using the ΔM images after they have been filtered, and using the last image as the reference volume since each previous image will contain a subset of data from this final image. On the subject of pre-processing, smoothing of arterial difference maps helped to improve fit stability but also had the effect of making vessels appear thicker compared to the difference images or maximum intensity projections. While this is not an issue for estimating inflow dynamics, it may give the appearance of higher blood volume that should not be mistaken for vascular remodelling. We found that the ability to resolve more distal vessels depended on the data quality and pre-processing choices, but we could not identify an optimal general approach. It is important to note that changes in the acquisition parameters will also influence the detectability of small vessels, and may require unique pre-processing strategies to achieve optimal results. In our case, we chose to implement an iterative Frangi filter to enhance ‘vesselness’, however several alternatives exist including the Jerman filter^36^ or total variation filter, among others.

Finally, it is important to note that for the native atlas projections, the average mATT for a specific ROI is weighted not only by the arrival time at the terminal ends of the macro-vasculature (i.e. before perfusing the tissue), but also by the mATT of blood that is passing through the region. This nuance must be considered when interpreting sources of variance while correlating mATT maps with other hemodynamic parameters.

### Future-work

From the perspective of more accurate fitting of mATT, it seems beneficial to more densely sample the signal response by adding more time-points. This could help to better characterize the saturation of the inflow signal, thereby improving the estimation of the breakpoint. Another consideration is that for very slow flowing blood, or in the case of long collateral pathway, the label may never arrive at downstream locations. Therefore, the measured inflow signal depends on the longest label duration selected in the scan and the label T1 decay. By increasing the number of time-points (at the cost of longer scan time) the presented model could be sensitized to smaller increases in mATT arising from possible vascular steal effects. It should also be noted that with higher acceleration factors there may come a trade-off in the capacity to delineate distal vessels (see preliminary results in figure S-4). However, this may not be a problem when the aim is to assess hemodynamics in the largest vessels, and advanced pre-processing using deep learning based techniques,^37^ or more customized data filtering may recover noisy signals at later time points.

Our method of projecting to the native atlas will enable prospective studies to correlate mATT with hemodynamic parameters such as CVR or parenchymal ATT. For instance, differences between macrovascular and tissue-level arrival times may serve as markers for subtle perfusion deficits at the level of small vessels. Additionally, a mismatch between ATT and mATT (as shown in figure 5) could indirectly indicate the status of leptomeningeal collaterals, with mATT providing a valuable reference point that conventional perfusion imaging lacks. Our examples highlight the heterogeneous nature of the disease in this patient group, as reflected by the large standard deviation in the rightmost panel of figure 3. With a sufficiently large sample size, it would be insightful to correlate mATT with specific disease markers to assess how much variation is driven by clinical versus structural parameters. For example, normalization of mATT through bypass surgery, compared to less-or non-affected contralateral vessels (as shown in figure 6), could indicate successful surgery, suggesting that rapidly arriving blood can adequately supply downstream tissue.

In summary, to fully assess the clinical relevance of the quantified mATT parameter, further steps are needed: (1) studies in healthy subjects for protocol optimization and comparison with ground truth modalities; (2) prospective case-control studies to better understand the effects of disease on mATT; and (3) longitudinal evaluations focusing on the impact of interventions and treatments.

## Conclusion

We introduce a novel model to quantify macrovascular blood arrival times based on 4D-MRA data. Moreover, we provide a method to represent sparse volumetric mATT values in a native atlas space, which facilitates spatial normalization and comparison with other three-dimensional hemodynamic parameter maps or anatomical structures of interest. The ability to quantify dynamic macrovascular information lays a foundation for future prospective studies seeking to understand the relationship between large-vessel and tissue-level hemodynamics, and monitor surgical treatment in steno-occlusive disease patients.

## Data Availability

All data produced in the present study may be made available upon reasonable request to the authors

## Acknowledgments

This work was supported by the W.M. De Hoop Foundation, the Janivo Foundation and Friends of UMC Utrecht & Wilhelmina Children’s Hospital, and an NWO VIDI grant awarded to A.A.B. (VI.Vidi.223.085). Thank you to Prof. David J. Mikulus for the insightful discussion.

