## Supplementary material for "A novel model to quantify blood transit time in cerebral arteries using ASL-based 4D magnetic resonance angiography with example clinical application in moyamoya disease": Suplementary

Supplementary Figures

Table S-1: Patient Data

| SUBJECT | Age | Sex | Diagnosis | Affected hemisphere  angiographically | Symptomatic  hemisphere | Previous  surgery |
| --- | --- | --- | --- | --- | --- | --- |
| subj1 | - | Female | Moyamoya disease | Both | None | Indirect left |
| subj2 | - | Female | Moyamoya disease | Both | Both | None |
| subj3 | - | Female | Moyamoya syndrome (Down syndrome) | Left | None | None |
| subj4 | - | Male | Carotid occlusion | Right | Right | Direct right |
| subj5 | - | Female | Moyamoya disease | Both | Right | Direct right |
| subj6 | - | Female | Moyamoya disease | Both | Left | Direct left |
| subj7 | - | Female | Moyamoya syndrome (Neurofibromatosis type I) | Both | Left | Indirect left and right |
| subj8 | - | Female | Moyamoya disease | Both | None | Direct left, indirect left/right |
| subj9 | - | Female | Moyamoya disease | Both | Left | None |
| subj10 | - | Male | Moyamoya disease | Both | Left | None |
| subj11 | - | Female | Moyamoya disease | Both | Left | None |
| subj12 | - | Male | Moyamoya disease | Both | Left | Direct and indirect left/right |
| subj13 | - | Male | Healthy subject | None | None | None |
| subj14 | - | Female | Healthy subject | None | None | None |
| Subjects were excluded due to artifacts in the 4D-MRA | | | |  |  |  |

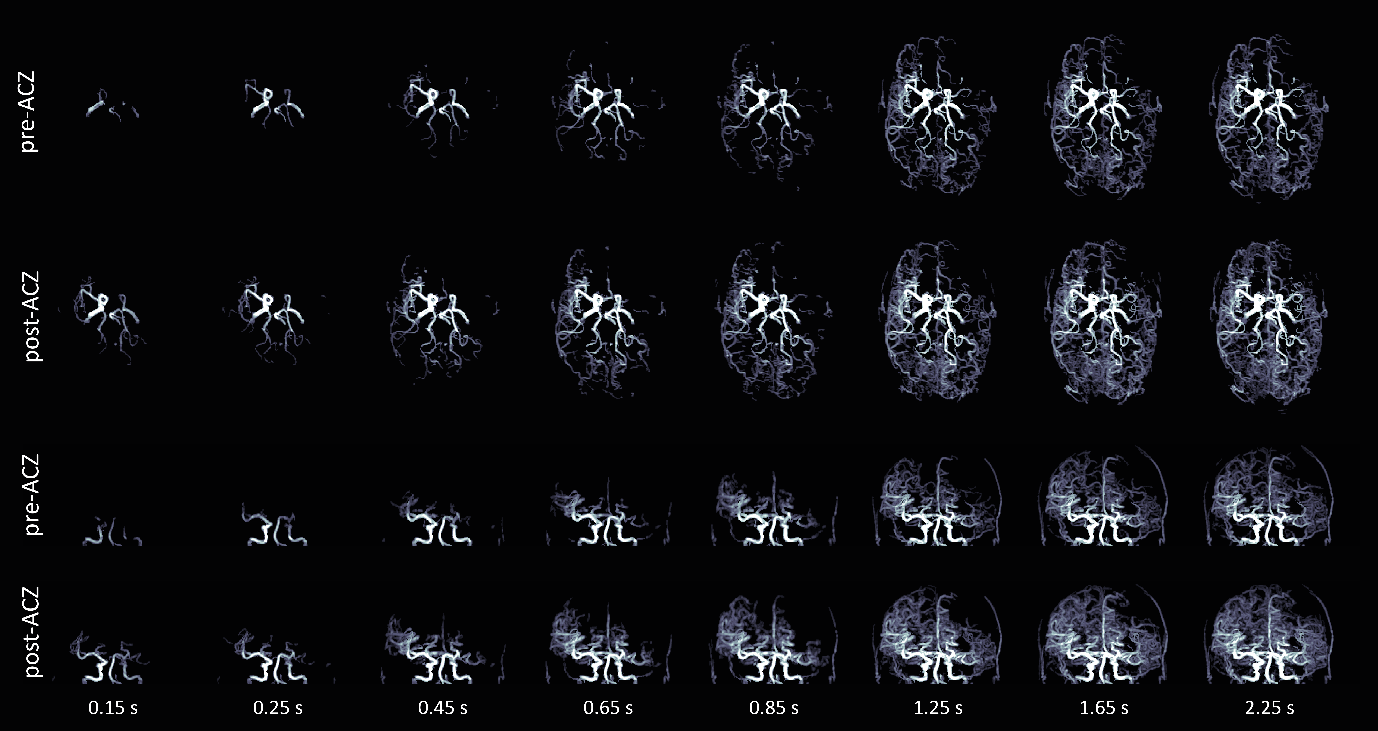

Figure S-1: The effect of Acetazolamide on the inflow signal. 4D-MRA images at progressive time-points both pre- and five minutes post-acetazolamide (ACZ) injection. Maximum intensity projections in the transverse (top) and coronal (bottom) orientation are shown.

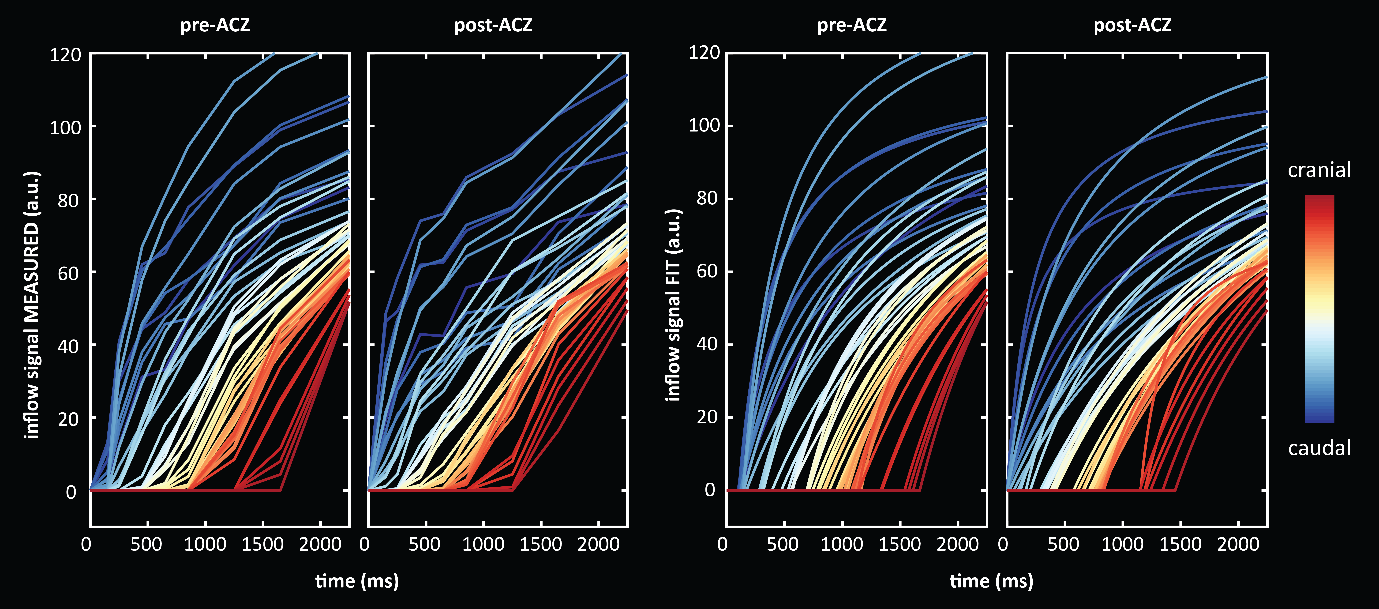

Figure S-2: measured (left) and fit (right) inflow curves (averaged per 3 slices) before and after ACZ injection in patient shown in figure 4 of manuscript and supplementary figure 1. Note the shortened arrival time.

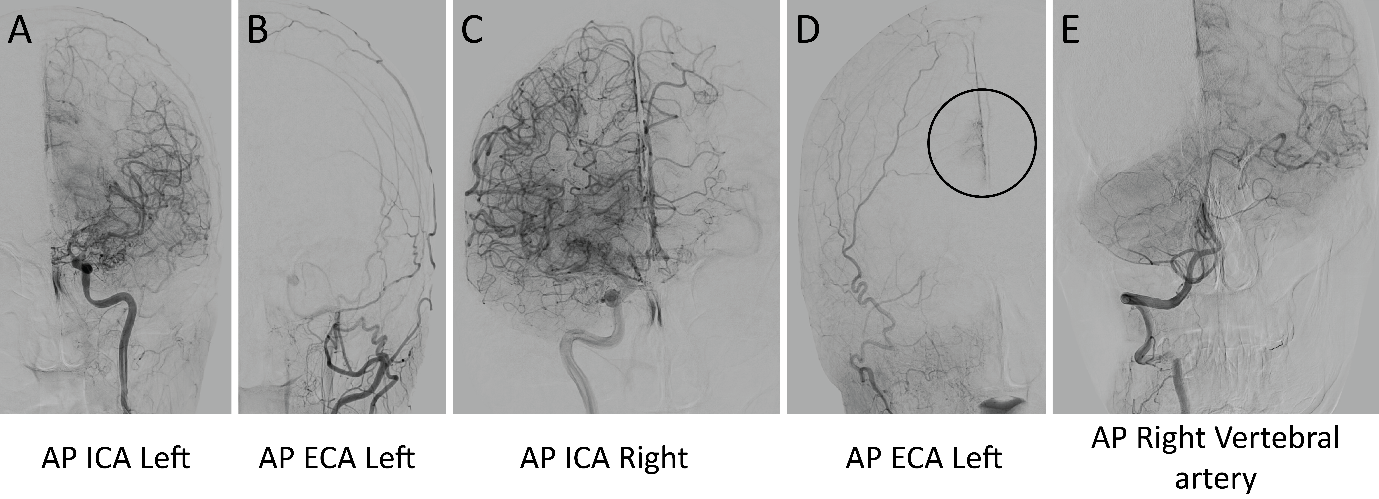

Figure S-3: Selected DSA images acquired at a referring hospital of the patient shown in figure 4 of the manuscript. A: AP image of the ICA on the left, showing occlusion of the distal ICA, the MCA and ACA cannot be identified, with extensive moyamoya collaterals; B: shows no naturally occuring ec-ic contribution from the left ECA; C shows the ICA on the right, with also extensive collateral formation around the distal ICA and no ACA identifiable. Contralateral flow of some bloodvessels; D: small contribution of the anterior territory form naturally occurring collaterals arriving from the ECA on the left side; E: contralateral filling of parts of the media territory on the left from the right vertebral artery.

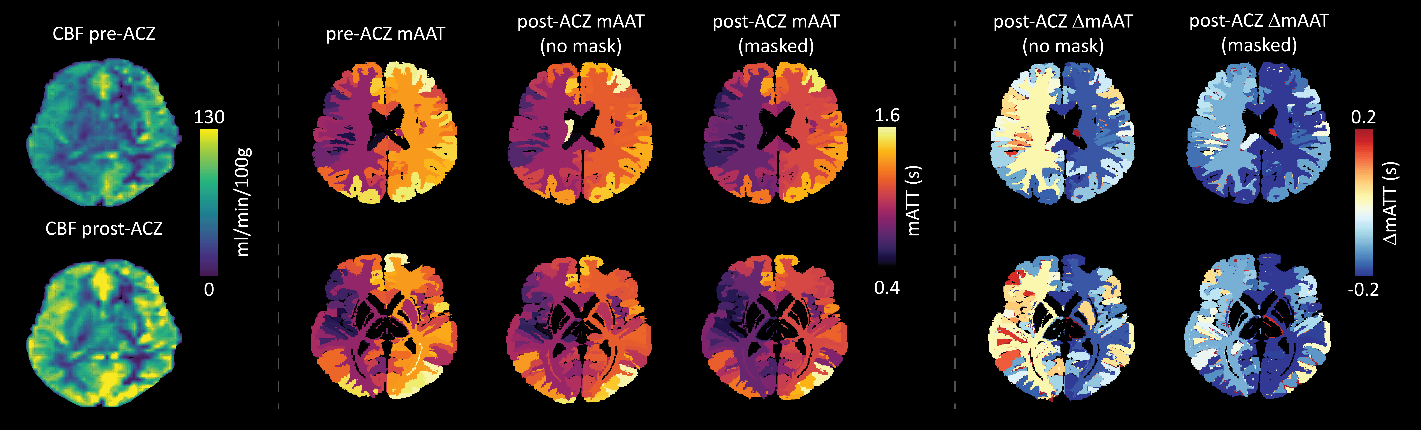
Figure S-4 – LEFT: Pre- and post-ACZ CBF maps in a patient shown in figure 5 of the manuscript. To investigate a possible source of the paradoxical mAAT values seen in the native atlas (contralateral to left-sided bypass), an additional analysis was performed in which the pre-ACZ vessel mask was used to mask the post-ACZ arrival time mask. After calculating the mean ROI values, the paradoxical arrival times on the unaffected side remained, however they became shorter than without the correction. This would suggest that the enhancement of distal vessels (and their associated longer mAAT) by the ACZ challenge can lead to increased arrival times in large regions where vessels with both short and long mAAT coincide. The observation that ΔATT in the contralateral hemisphere shows less decrease after ACZ injection may indicate a stronger contribution from leptomeningeal collaterals.

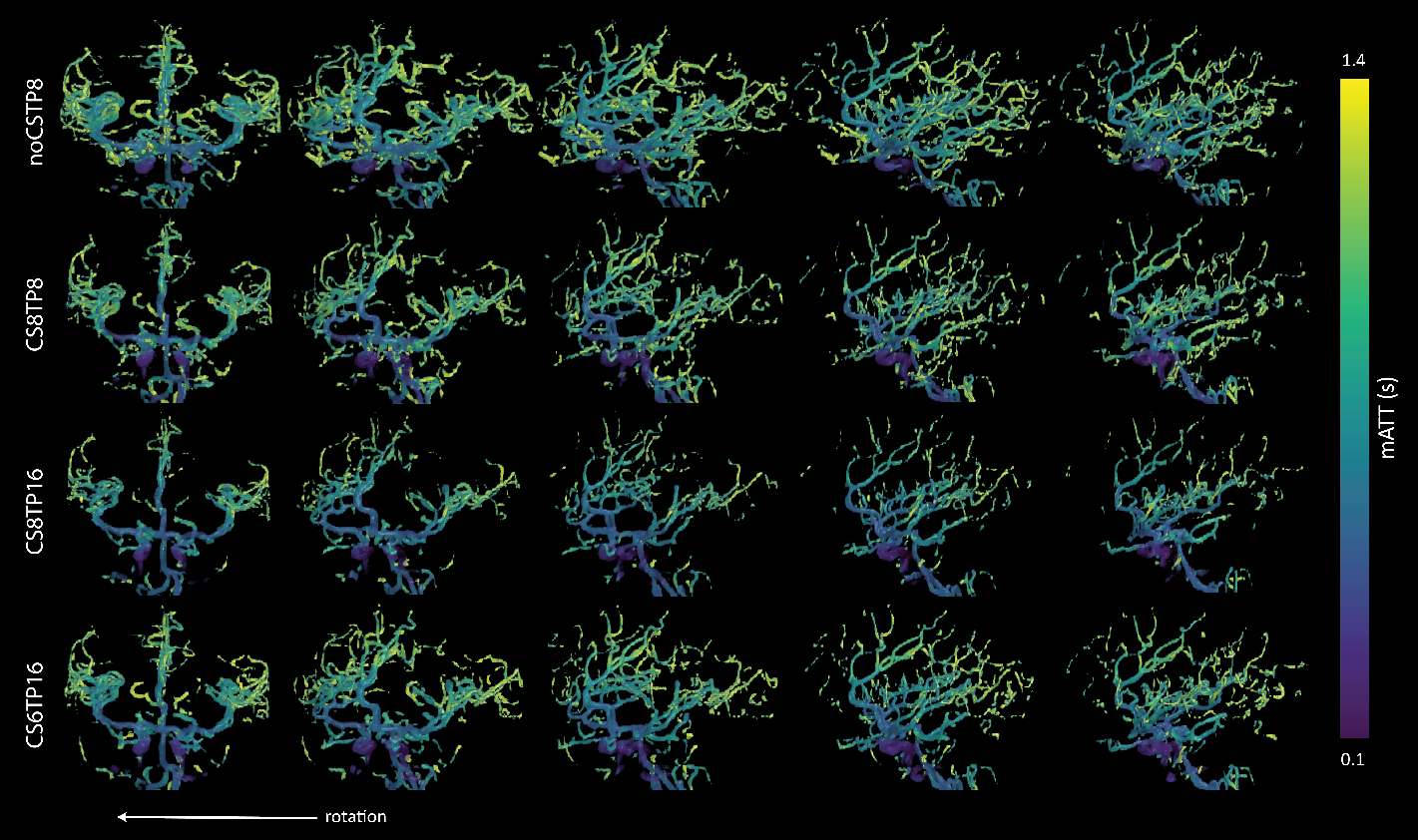

Figure S-5: The effect of increasing the number of time-points and accelerating the data acquisition using two different compressed sense (CS) factors. Increasing CS, can reduce scan time, but it comes at the cost of SNR at the distal vessels. A prospective study in healthy control subjects is warranted to determine the optimal balance of acceleration (to acquire more time points) and improved accuracy of the mAAT fit. The ‘noCSTP8’ scan (first row) was a reference scan with the same parameters as described in the main manuscript: 3D GRE readout, 8 time-points at 100, 200, 400, 600, 800, 1200, 1600 and 2200 ms, 75% keyhole central size, turbo factor=60, SENSE factor=3x1.5 (right-left, feet-head direction), half scan (partial Fourier) factor = 0.8×0.8 (right-left, feet-head direction), TR/TE/flip angle = 6 ms/1.97 ms/11°, acquired voxel size 1×1.4×1.6 mm^3^, reconstructed voxel size 0.6×0.6×0.8 mm^3^, field-of-view = 200×200×120 mm^3^, scan time: 6 min 16 s). For the images shown second row, a compressed sense factor of 8 was used leading to an approximate total scan time of 4 minutes. For the third row, a compressed sense factor of 8 was used and the number of time-points were doubled to 16 ( 100 200 300 400 500 600 700 800 900 1000 1200 1400 1600 1800 2000 2200) leading to an approcimate total scan time of 8 minutes. In the fourth row, the compressed sense factor was reduced to 6 using the same 16 time-points leading to an approximate scan time of 11 minutes and 20 seconds.

- MATLAB code for fitting function will be made available upon publication
